# WHAT IS THE RISK OF DEVELOPING A SEVERE FORM OF COVID-19 INFECTION AMONG ADULTS WHO CURRENTLY SMOKE COMPARED TO EX-SMOKERS? A protocol for systematic review and meta-analysis

**DOI:** 10.1101/2022.11.10.22282181

**Authors:** Taagbara Jolly Abaate, Abueh Nukoamene Prince

**Affiliations:** Public Health/Community Medicine Department, University of Port Harcourt Teaching Hospital, Rivers state, Nigeria

**Keywords:** COVID-19, Adults, Ex-Smokers, Severity, Smoke, Humans, Smoking

## Abstract

**Introduction:** Smoking is one of the lifestyle choices associated with an increased risk of chronic health conditions and poorer COVID-19 outcomes. Because it is known that the lungs recover after quitting smoking, a direct comparison of the severity of COVID-19 infection in current and former smokers needs to be investigated.

**Methods and analysis:** The Preferred Reporting Items for Systematic Reviews and Meta-Analyses Protocol (PRISMA-P) 2015 Checklist was used. Non-randomized studies will be searched in PubMed, Cochrane CENTRAL library, Embase, and Epistemonikos from December 2019 to the present. Hand-searching of grey literature, key journals, and reference lists will be conducted

This review will include studies of current and former smokers, with the main outcome being ICU admission, assisted respiration, or death. Two independent reviewers will select primary studies and abstract data from them. The Newcastle-Ottawa checklist will be used to assess the risk of bias, and the Grading of Recommendations Assessment, Development, and Evaluation (GRADE) framework will be used to assess the quality of cumulative evidence. RevMan 5.4 will be used for data analysis.

The I^2^ statistic will be used to evaluate heterogeneity. For similar studies, the fixed-effect method of a meta-analysis will be used; otherwise, a random-effect model will be used. The qualitative synthesis will be used for studies that are ineligible for the quantitative approach.

**Ethical consideration and dissemination:** Because published data will be reviewed, no ethical approval is required. Our findings will be presented at national and/or international conferences, and they will be published in a peer-reviewed journal.

**PROSPERO registration number CRD42022368552:** 

**ARTICLE SUMMARY:** 

**Strengths and limitations of this study:**

1. This is a focused research question comparing the current and ex-smokers risk of contracting the severe form of COVID-19.
2. This systematic review and meta-analysis will provide evidence of the dangers of smoking during the COVID-19 pandemic.
3. The PRISMA-P reporting guidelines were strictly followed while writing this protocol.
4. Study selection will be carried out by two independent reviewers and a third person will intervene if a disagreement arises.
5. A potential limitation is that an observational study design will be used in this systematic review.

## INTRODUCTION

The pathogen responsible for the coronavirus disease that began in the Chinese province of Hubei in 2019 (COVID-19) is the Severe Acute Respiratory Syndrome Coronavirus (SARS-CoV-2). (1,2)

COVID-19 unexpectedly posed a significant risk of international spread, a significant risk of trade restrictions, and had a serious public health impact;(3) thus, it was designated as a ‘Public Health Emergency of International Concern (PHEIC)’ by the World Health Organization (WHO). (4) The current COVID-19 outbreak is one of six PHEICs declared by WHO thus far, the others being the 2009 novel flu pandemic, wild polio in 2014, West African Ebola in 2014, Zika virus in 2016, and the 2018 Ebola outbreak. (4) As a result, on March 11, 2020, the WHO declared COVID-19 a pandemic, (5) and it has spread to almost every country in the world, impeding the growth of these countries where novel coronavirus cases have been reported. (6)

Risk factors for COVID-19 include demographic elements such as age and sex, as well as dietary and lifestyle choices, underlying illnesses, and laboratory findings. (7) Most people who develop COVID-19 will experience mild-to-moderate illness primarily affecting the respiratory system and recover at home. (8) In more severe cases, patients may require specialized care (e.g., admission to hospital or intensive care unit (ICU) and assisted ventilation) as the disease can progress to respiratory failure, affect multiple organ systems, and eventually death. (9)

Smoking is one of the lifestyle choices known to increase the risk of chronic health conditions linked to poorer COVID-19 outcomes. (18) Cigarette smoking is the single most preventable lifestyle cause of death worldwide, and it has been linked to a plethora of illnesses including respiratory diseases. (11-13) As a result, strict public health measures must be implemented in every country and at every level to ramp up campaigns to prevent initiation and support quitting. (10) Smoking may raise the chance of systemic infections (11) by altering the function of the cellular and humoral immune systems. (12) Smoking reduces the function of polymorphonuclear leukocytes, which play an important role in the host’s defense against bacterial infection. (12) Cigarette smoke impairs respiratory immune defense by inducing peri-bronchiolar inflammation and fibrosis, impairing mucociliary clearance, and disrupting the respiratory epithelium. (13)

In the COVID-19 pandemic, concerns have been raised about the clinical outcomes for smokers, whether they are equally susceptible to infection, and whether nicotine has any biological effect on the SAR-CoV-2 virus. (14) The SARS-CoV-2 virus enters epithelial cells via the angiotensin-converting enzyme 2 (ACE-2) receptor2, (15) and studies show that current smokers’ airway and oral epithelium exhibit increased gene expression,(13,16) which may increase their risk of contracting SARS-CoV-2. (17) COVID-19 virus’s ability to infect T cells through the angiotensin-converting enzyme 2 (ACE2) receptors and a cluster of differentiation 147-spike protein has also been linked to lymphopenia,(18,19) a marker of severity in patients infected. (20–22)

Other studies have posited that nicotine may have an inhibitory effect on ACE-2 receptors,(23) but despite these uncertainties, both former and current smoking status is known to increase the risk of respiratory viral and bacterial infections,(24) as well as worse outcomes once infected. (25)

Furthermore, it has been demonstrated that the risk of infection caused by smoking is reduced by half and eventually disappears after smoking cessation. (26)

According to a recent review,(27), there is limited evidence that disease severity in those hospitalized for COVID-19 is greater in current or former smokers than in never smokers. Also, there is insufficient evidence to conclude that infection, hospitalization, or mortality are increased in current and former smokers. (28) Studies that have been done to date to evaluate the smoking status and the negative progression of COVID-19 have typically involved very few cases, and confounding variables may prevent the effect of smoking from being distinguished. (29)

### RATIONALE FOR THIS SYSTEMATIC REVIEW AND META-ANALYSIS

The evidence for smoking and COVID-19 risk is less clear. Potential misclassification of current smokers in empirical studies included in a previous meta-analysis understates the impact of current smoking on the risk of severity and mortality in this patient population. (16) It is known that the lungs recover after someone stops smoking,(30) including former smokers in the exposed group may bias the effect estimate to the null.

To date, there has not been a direct comparison between current and ex/former smokers and the severity or negative progression of COVID-19 infection. Several studies have found that smokers are more likely than nonsmokers to develop severe COVID-19 respiratory disease(27). Understanding these issues is critical for assessing clinical risk, developing clear public health risk communication, and identifying intervention targets.

Previous studies were mostly done in China and conducted in 2020, and as more evidence evolves on the ongoing pandemic, there is a need for focused research in this area.

Again, one of WHO’s global targets for non-communicable diseases in 2025 is to have a 30% relative reduction in the prevalence of current tobacco use in adults and the findings from this study will buttress the global campaign against smoking. (31)

### OBJECTIVES OF THE REVIEW

1. To determine the risk of severe disease (defined as; admission to ICU, mechanical ventilation, and death) in current smokers compared to ex-smokers.
2. Effect of comorbidities on the outcome of ex-smokers and current smokers with the negative progression of COVID-19.

## METHODS

This proposed systematic review and meta-analysis protocol conforms with the Preferred Reporting Items for Systematic Reviews and Meta-Analyses Protocol (PRISMA-P) 2015 Checklist (32) and other protocols. (33,34) This protocol is registered with PROSPERO 2022.

### Eligibility criteria (Inclusion/exclusion)

This systematic review will include studies published in any language on current and former smokers over the age of 18 diagnosed with covid-19 using the gold standard Polymerase Chain Reaction (RT-PCR) test. (35)

### The PICOs strategy for inclusion criteria

Population/participant: Adult smokers18 years and older

Intervention/exposure: Smoking (current smokers)

Comparator: Former smokers

Outcome: The severity of COVID-19 infection is measured as admission to the Intensive Care Unit (ICU) or the High Dependency Unit (HDU), mechanically ventilated, or death. (16)

Study design: To answer the research question, observational studies (retrospective, prospective clinical studies, case series, and descriptive studies) published from December 2019 to date will be included because cases of COVID-19 were first reported in December 2019. (1)

### Exclusion criteria

Systematic reviews, opinion pieces, non-clinical studies, studies that do not report data on current and ex-smokers, studies that report on the pediatric age group, and studies examining other coronaviruses will be excluded. The PRISMA flow diagram will show excluded papers.

### Patient and public involvement

This is a systematic review protocol, therefore, individual patient data will not be presented. An extensive literature search will be carried out from defined databases. For this reason, no patient will be involved in the study planning and application process during either the analysis or dissemination of results.

### Information sources

An electronic search for relevant articles will be conducted by the lead author (AJT) across PubMed, CINAHAL, Embase, Web of Science, Cochrane CENTRAL, Scopus, and Epistemonikos from December 2019 to the present.

### Other sources

#### Grey literature

Grey literature will be accessed via the British national bibliography for report literature, science citation index for conference abstracts, OpenGrey database, and ProQuest Dissertations.

#### References list of included studies

Hand-searching of reference lists will be performed for additional relevant studies that have been missed from a database search to minimize publication bias.

#### Searching of key journals

This will include journals such as; SAGE, AJOL, and the Lancet through the web of science citation index and hand searching.

### Search strategy

For a comprehensive search, search terms will include keywords and controlled vocabularies and these will be combined using Boolean operators. Wildcards or truncation will be applied to search for words ending in different forms. A full search strategy is attached however, here is an example of some of the search terms; COVID-19 OR Sars-CoV-2 OR Coronavirus infection AND severity OR progression OR (admission to ICU) OR mechanical ventilation OR death AND (smoking status) OR (current smoker) OR smok* OR (cigarette smoking) OR (tobacco smoking) AND Human OR (adult 18 years and older) AND observational studies OR (prospective studies) OR (cohort studies) OR (case-control studies) OR (descriptive studies). The search strategy developed will be adapted to all databases. There will be no language restrictions and articles published from December 2019 when COVID-19 was discovered in China to the present will be searched for. The PRISMA flow diagram will show the sources of information and articles retrieved.

### Data management

Articles identified through various means will be exported into the ZOTERO reference manager for duplicate checking and retrieval. This will be followed by a screening process in which forms will be created and tested. A standardized form created solely and piloted for this research will be used for data extraction. The form will be adapted as needed and the results will be entered into software for analysis. Meta-analysis will be performed if we find sufficient studies that are homogeneous.

### Selection process

The eligibility screening will take place in three stages: title, abstract, and full report. Two reviewers (AJT & ANP) will independently screen the articles for eligibility and unrelated studies eliminated at the title stage. Potential studies for inclusion based on abstract and full text would be read and any disagreements will be resolved through discussion. We will attempt to group similar studies.

### Data collection process

Data collection forms will be developed and piloted in three or four studies. Written instructions on what data to collect will be given to the two reviewers to follow. The lead reviewer (ATJ) and a second reviewer (ANP) will independently carry out the data abstraction process. The summaries will be compared, and any disagreements will be resolved by convening a meeting for discussion. If an agreement cannot be reached, a third reviewer will be contacted for a final decision.

### Data item for collection

The following details will be extracted from the included articles: participants’ age, sex, socioeconomic status, and comorbidities; research details (author, objectives of study, journal, date, country, study design, study period, sample size, statistical analysis, level of evidence, and financing). Exposures include present and past smoking habits, as well as clinical outcomes like admission to the ICU or HDU, the need for mechanical ventilation, or death. Results will contain participant summary statistics e.g. risk ratios. Additionally, each report’s ID, date, and name of the data extractors will be recorded.

### Addressing missing data

If any of the selected articles lacks information, we will contact the corresponding author and the principal investigator via email and phone to obtain clarification and the missing data. If this is not possible, the data will be deleted and discussed in the Discussion section.

### Outcomes and prioritization

The main outcome of this systematic review and meta-analysis will be studies that reported summary statistics on the progression or severity of COVID-19 infection in current and former smokers that were admitted to the hospital. Severe COVID-19 is defined by acute respiratory distress syndrome, partial pressure of oxygen (SPO_2_) less than 94% requiring mechanical ventilation or resuscitation, intensive care unit admission, and death. (16) another outcome measure that will be sought include the effect of comorbidities on the outcome of COVID-19 infection in ex-smokers and current smokers.

### Risk of bias (quality) assessment

The Newcastle-Ottawa Scale checklist will be used to evaluate the quality of the studies considered for the analysis at both the outcome and study levels. The risk of bias in non-randomized trials and observational studies is assessed using three subscales on the checklist: selection, comparability, and outcome.

### Assessment of Heterogeneity

Forest plots will be created to show both the pooled odd ratio (OR) estimates and the study-specific OR estimates with their confidence intervals. Heterogeneity will be evaluated statistically using the Chi-squared test of heterogeneity (Mantel-Haenszel or Cochran Q test with P value > OR 0.05) as well as visually in the forest plot for overlapping confidence intervals and the summary effect. I^2^ statistic, (31) (the percentage of observed variation attributable to heterogeneity), meta-regression, and tau (the actual amount of observed heterogeneity) will be used to address the heterogeneity. Significant heterogeneity is defined as I^2^ greater than 50%, and low heterogeneity is defined as I^2^ less than 50%.

### Data synthesis

Both qualitative (text word and tabulation) and quantitative analysis will be performed. An odd ratio with 95% confidence intervals will be extracted from each study as the effect measure for dichotomous outcomes. Where the studies are sufficiently similar and the I^2^ is less than 50% (36), a fixed effect model will be used for the quantitative analysis. However, the random effects meta-analysis method will be used to pool the effect measure in cases where heterogeneity is present (I^2^>50%). (37) To assess the robustness of our findings, sensitivity analyses will be conducted on studies not eligible for meta-analysis or by removing a study (based on sample size or one that scored lower on the Newcastle Ottawa scale) to see if the effect estimate changes.

Studies from China where COVID-19 was first reported would be subjected to subgroup analysis, as would studies with similar study designs and studies from the rest of the world. The lead reviewer would use Review Manager Version 5.4 software to carry out all statistical analyses.

### Assessment of meta-biases

If there are enough studies, selective reporting of non-significant results or publication bias will be assessed using a funnel plot.

### Confidence in commutative evidence

To assess the evidence, the Grading of Recommendations Assessment, Development, and Evaluation (GRADE) framework will be used. (38) This will consist of five steps, with the final quality of evidence being presented as ‘high, “moderate,’ ‘low,’ very low.’ These guidelines state that observational studies will be given an a priori rating of low quality and will then be either upgraded or downgraded.

Large effects (when the effect is so large that bias common to observational studies cannot possibly account for the result), dose-response relationships (when the result is proportional to the level of exposure), and situations in which all potential confounders would only reduce the observed effect, making it likely that the actual effect is larger than the data suggests are reasons to upgrade the certainty of the evidence. (39,40) Reasons to downgrade the certainty of the evidence include the risk of bias, inconsistencies between studies, imprecision, indirectness of the evidence, and publication bias. (41, 42)

## DISCUSSION

The quality of evidence found in this meta-analysis will be thoroughly discussed in light of what is known and what this study adds. The limitations of included studies, the strengths and weaknesses of the review methods, and finally, a conclusion on findings and the implications for current practice and future research will be stated.

## DISSEMINATION

Our results will be presented at national and/or international conferences and published in a peer-reviewed journal. Any changes made to the protocol during the review process will be documented in the manuscript.

## Data Availability

All data produced in the present study are available upon reasonable request to the authors

## ETHICAL CONSIDERATIONS

This systematic review and meta-analysis will compile previously published research, in which participants would have given their consent voluntarily and knowingly. This removes the requirement for ethical approval.

## ACKNOWLEDGMENT

I would like to express my heartfelt appreciation to Dr. Deswosu Ihuoma, who critically reviewed the study protocol and restructured a paragraph in this work’s background section.

## AUTHOR CONTRIBUTIONS

AJT conceptualized and wrote the methodology as well as the first draft of this protocol for a systematic review and meta-analysis, even though all authors contributed to its development.

## PATIENT AND PUBLIC INVOLVEMENT

Patients and the general public were not involved in the research’s planning, design, execution, or reporting. PATIENT CONSENT FOR PUBLICATION

Not necessary.

## PROVENANCE AND PEER REVIEW

Not commissioned; externally peer-reviewed.

## OPEN ACCESS

This open-access article is distributed under the Creative Commons Attribution Noncommercial (CC BY-NC 4.0) international license, which enables others to distribute, remix, adapt, and build upon this work noncommercially and license their derivative works under various conditions, provided that the original work is properly cited, the proper credit is given, any changes made are indicated, and the use is for noncommercial purposes.

See http://creativecommons.org/licenses/by-nc/4.0/.

